# Breaking the seasonal barrier: feasibility of cuffless fingertip-based continuous blood pressure monitoring in older adults during winter exercise

**DOI:** 10.64898/2026.04.14.26350440

**Authors:** Natsuko Mizutani, Satoshi Nishizawa, Yukie Enomoto, Hiroteru Okamoto, Ryushin Baba, Ayaka Misawa, Kahimi Takahashi, Yuina Tada, Yu-Ching Lin, Wen-Pin Shih

## Abstract

While the need for continuous blood pressure (BP) monitoring in Japan is high, there are no commercially available cuffless devices for personal daily monitoring use. Fingertip-based sensors are a promising alternative as they eliminate the discomfort of repeated cuff inflation. However, their reliability during winter has been a major technical limitation due to cold-induced peripheral vasoconstriction. This study aimed to address this issue by validating a novel fingertip-based continuous BP monitor used by exercising adults during summer and winter. Eleven community-dwelling older adults (mean age, 73.1 ± 8.8 years) were included in this seasonal comparative study. During exercise, we compared a personal fingertip-based continuous monitor (ArteVu) with a standard oscillometric cuff device (Omron) in summer (mean, 26.5°C) and winter (mean, 7.4°C). The study also evaluated the device’s accuracy during exercise-induced BP fluctuations and seasonal environmental changes. Awareness of the participants regarding BP management was also assessed using questionnaires. There were strong correlations for systolic BP (SBP) between summer and winter (r = 0.93 in summer; r = 0.88 in winter). Although the mean difference for the SBP was higher in winter than in summer (3.1 ± 11.2 mmHg vs. 0.2 ± 9.4 mmHg), the values remained within a clinically acceptable range for personal monitoring. Notably, 72.7% of participants reported that the ease of using the fingertip-based device significantly increased their awareness and motivation for daily BP management. This study confirms the feasibility of cuffless fingertip-based continuous BP monitoring across different seasons, including in winter. By overcoming the seasonal limitations, this device fills a critical gap in the Japanese health-monitoring market. Our findings support the development of smaller and more portable models, representing a shift from traditional “snapshot” cuff measurements to continuous and integrated lifestyle monitoring for older adults.

## Introduction

### The current gap in the Japanese market

Although hypertension affects millions of individuals in Japan, the tools available for daily blood pressure (BP) management remain largely unchanged. Although accurate, conventional oscillometric cuff devices only provide “snapshot” measurements and are cumbersome as they require arm exposure and cuff inflation, which often result in poor adherence. Despite the clinical need for continuous BP monitoring to capture dynamic fluctuations, continuous BP monitors designed for personal and everyday are commercially unavailable in Japan [1][2][3] [4][5][6].

### The promise of fingertip-based continuous monitoring

A novel fingertip-based mechanical sensor (ArteVu, US FDA K244007) that is cuffless and allows for continuous and non-invasive BP monitoring was developed to address the clinical need. By simply placing a finger on the sensor, users can obtain their BP in real-time without the discomfort of repeated cuff inflation [4]. This device may potentially shift BP management from a traditionally discrete clinical task to a continuous and integrated part of a healthy lifestyle.

### The “winter challenge”: why this study is critical

The primary technical concern for any fingertip-based BP sensor is the influence of the environmental temperature. During winter, peripheral vasoconstriction significantly reduces blood flow to the fingertips, which may compromise signal detection [5]. Therefore, if a fingertip device cannot perform reliably in winter, its utility as a personal monitoring tool is severely limited. After our preliminary validation of the device’s accuracy (Mizutani et al., 2025[6]), repeating the validation in real-world conditions was warranted.

This study validated the accuracy of a fingertip-based continuous BP monitor when used by exercising adults during summer (approx. 26°C) and winter (approx. 7°C). By comparing the measurements between seasons, we aimed to:

1. Demonstrate that the fingertip sensor can reliably capture BP signals even during winter.
2. Validate the clinical feasibility of a continuous cuffless BP monitoring device as a superior alternative to traditional cuff measurements for enhancing long-term health awareness.

## Materials and Methods

### Study Design and Participants

This seasonal comparative study enrolled 11 community-dwelling older adults (mean age 73.1 ± 8.8 years; 4 men, 7 women) who had participated in a health promotion program. The purpose of this study was to assess the performance of a novel fingertip-based continuous BP monitor under different outdoor temperatures. All participants provided written informed consent. This study was approved by the Ethics Committee of Kyorin University (Approval No. 2024-28 and 2024-72).

Note that this study actually began on July 13, 2024, and the data used for analysis was extracted from the period ending July 20, 2025. In addition to holding this event, we explained the details to the organizers in advance and obtained their permission to hold it on the day of the event. At each session, the principal investigator personally explained the study details in person, and only participants who gave their consent were allowed to participate; consent forms were signed. It should be noted that there were no minors among the participants in any of the review meetings. As a further note, for the purpose of writing this paper, I reviewed the recorded data sheets stored in a locked cabinet in the laboratory from March 1 to 20, 2025. Although the participant data had been compiled in an anonymized form, it was still possible to identify the participants.

### Measurement devices (Fig 1)

1. **Fingertip-based continuous BP monitor (ArteVu**, San Jose, California): A mechanical sensor-based device that detects BP signals from the fingertip. Unlike optical PPG sensors, it utilizes a mechanical mechanism to detect arterial pulsations. For each participant, initial calibration was performed using a standard cuff-based measurement at the start of each session.
2. **Reference device (Omron HCR-7204T, Japan):** A validated and automated oscillometric upper-arm cuff BP monitor was used as the gold standard for comparison [8].

**Figure 1.**
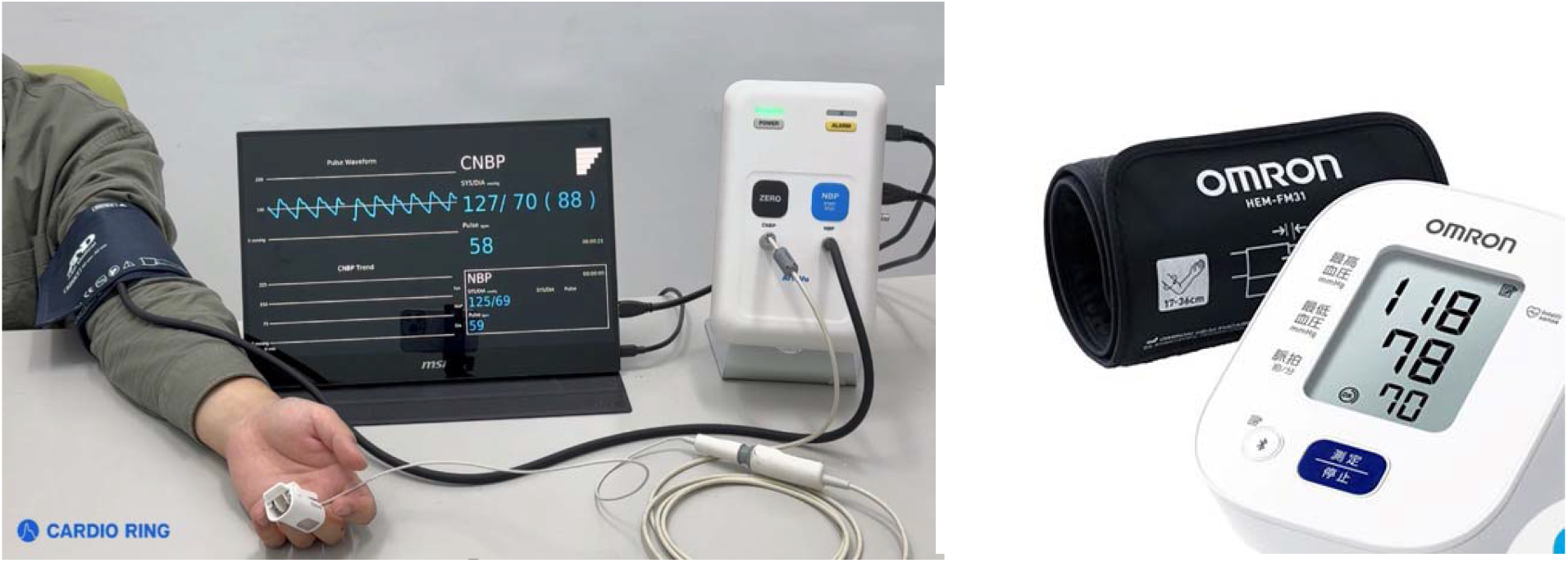
Blood Pressure Monitoring Devices. The ArteVu finger-tip blood pressure monitor (left photo), developed by Cardio Ring Technologies, San Jose, California detects signals using a mechanical mechanism applied to the fingertip. The photo on the left shows the first medical model, which includes a cuff because it requires initial calibration. For this comparison, we used the HCR-7204T model from Omron (right photo), a leading manufacturer of home blood pressure monitors.

**Figure. 2.**
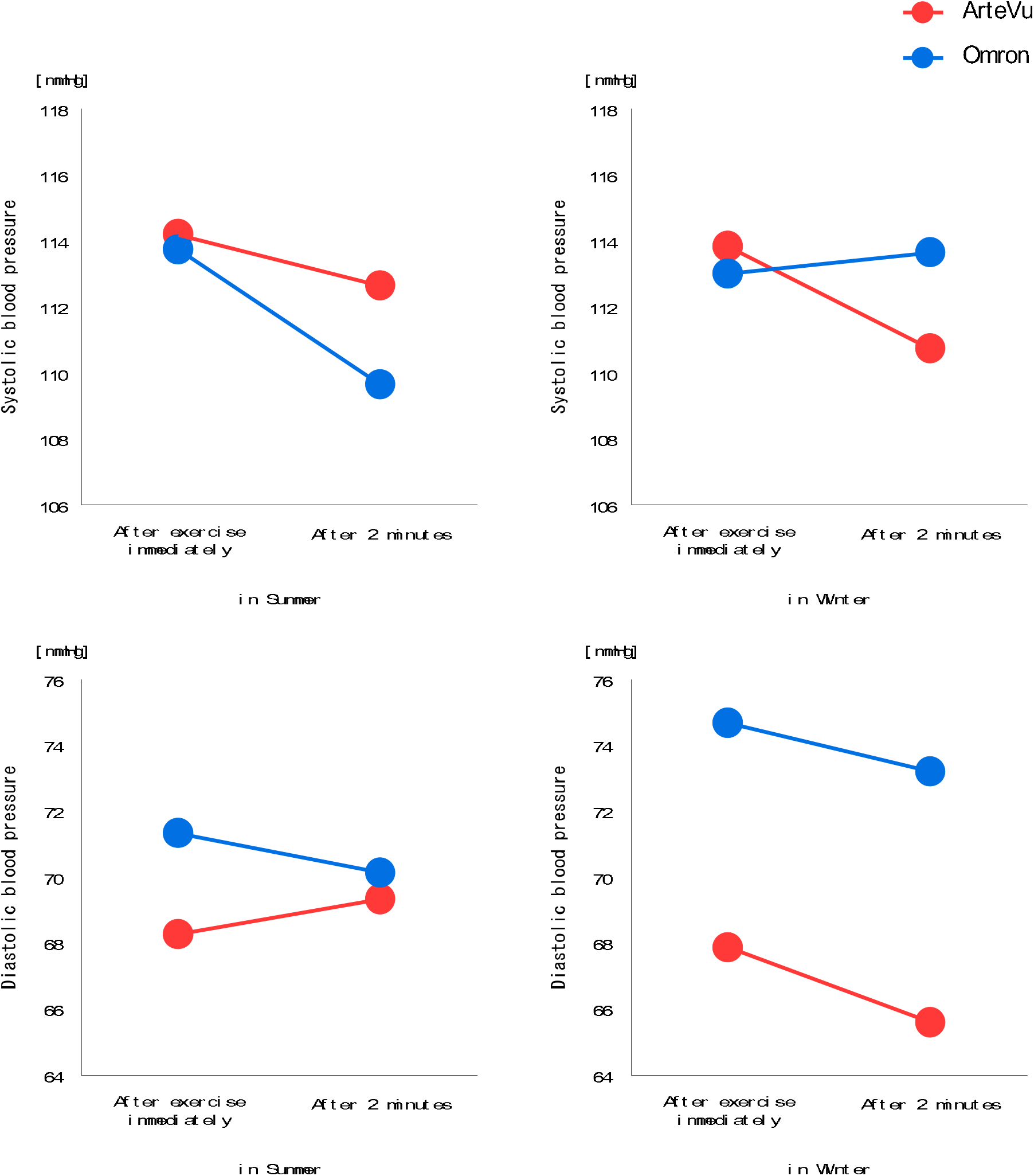
Difference of mean in systolic and diastolic blood pressure of each equipment (ArteVu : red, Omron : blue), between seasons or times after exercise. Through 3-way ANOVA, only equipment factor in diastolic blood pressure showed significant difference. No significant interaction was shown.

### Exercise protocols

- **Solar-Pole Exercise:** Participants performed stretching exercises. They were also provided a walking program using specialized poles to enhance posture and core stability, along with light aerobic exercise set to music.
- **Health and Wellness Classes:** A combination of light strength training and stretching exercises.

Each session lasted approximately 60–90 minutes, and the room temperature was maintained at 22°C–25°C for both sessions.

### Data Collection Procedure

BP measurements were obtained at three time points in both summer and winter:

1. **Rest:** After sitting quietly for at least 5 minutes before exercise.
2. **Immediately Post-Exercise:** Within 1 minute after completing the exercise program to obtain the peak BP response.
3. **Recovery:** Two minutes after cessation of the exercise. At each time point, measurements were obtained simultaneously or in immediate succession using both the fingertip-based monitor and Omron cuff device. For the fingertip monitor, participants were instructed to maintain their hand at heart level to minimize errors caused by hydrostatic pressure.

### Questionnaire on BP Awareness

After completing the measurements in both seasons, the participants completed a structured questionnaire. The questions focused on their previous BP monitoring habits, perceived ease of use of the fingertip device compared to traditional cuffs, and whether using the continuous monitor increased their awareness of personal BP management.

## Statistical Analysis

Continuous variables are presented as mean ± standard deviation (SD). The correlation between fingertip- and cuff-based measurements was evaluated using Pearson’s correlation coefficient (r). The agreement between the two modalities was evaluated using Bland–Altman plots, calculating the mean difference (bias) and limits of agreement (±1.96 SD). Data from summer and winter were evaluated separately to identify seasonal impacts on device performance. Statistical significance was set at p < 0.05.

## Results

### Participant characteristics

Table 1 presents the characteristics of the 11 participants (7 women, 4 men; mean age, 73.1 ± 8.8 years). The group included individuals who had different BP monitoring habits, ranging from daily measurement to almost never. Notably, several participants (ID: S3, H1, H2, H5, and H6) had previously detected elevated BP, making them an ideal demographic for evaluating a novel personal monitoring tool.

**Table 1.**
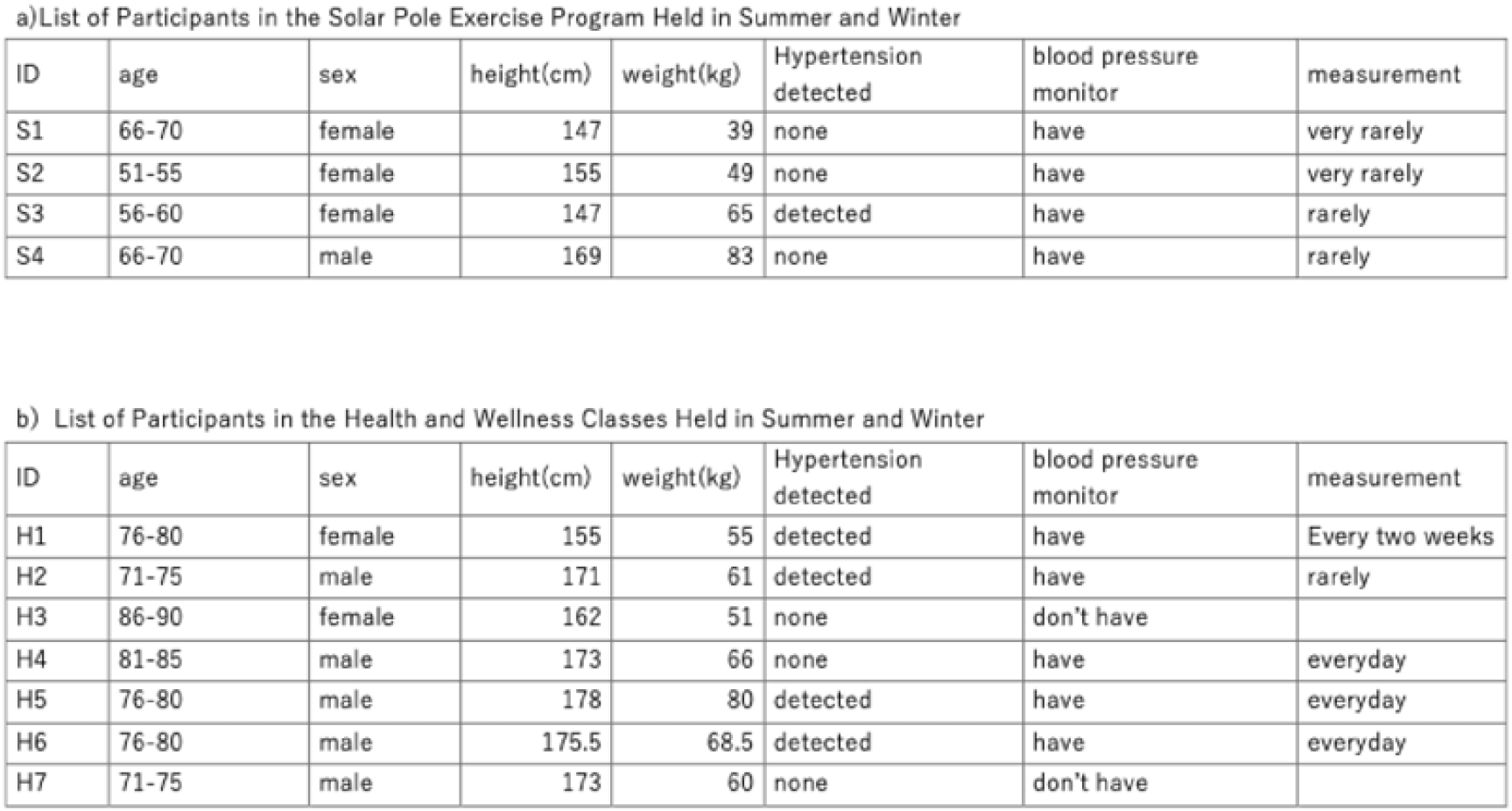
Participants.

### Device performance across seasons and exercise states

Table 2 presents the BP readings at the three time points for both summer and winter. The fingertip sensor demonstrated a higher degree of responsiveness to BP changes compared to the reference device.

**Table 2.** Blood pressure readings before exercise, immediately after exercise, and 2 minutes later in different seasons.

a) Solar Pole Exercise
| Summer | After exercise immediately |  |  |  | After 2 minutes |  |  |  |
| --- | --- | --- | --- | --- | --- | --- | --- | --- |
|  | Omron |  | ArteVu |  | Omron |  | ArteVu |  |
| S1 | 100 | 59 | 96 | 57 | 101 | 59 | 96 | 51 |
| S2 | 92 | 62 | 92 | 54 | 90 | 59 | 77 | 56 |
| S3 | 94 | 65 | 101 | 63 | 94 | 67 | 102 | 62 |
| S4 | 111 | 77 | 114 | 80 | 112 | 82 | 118 | 80 |

| Winter<br>ID | After exercise immediately |  |  |  | After 2 minutes |  |  |  |
| --- | --- | --- | --- | --- | --- | --- | --- | --- |
|  | Omron |  | ArteVu |  | Omron |  | ArteVu |  |
| S1 | 98 | 59 | 94 | 57 | 93 | 60 | 94 | 53 |
| S2 | 97 | 69 | 99 | 54 | 97 | 64 | 88 | 51 |
| S3 | 108 | 71 | 102 | 60 | 94 | 68 | 94 | 56 |
| S4 | 131 | 92 | 133 | 76 | 143 | 96 | 132 | 82 |

b) Health and Wellness Classes
| Summer<br>ID | After exercise |  |  |  | After 2 minutes |  |  |  |
| --- | --- | --- | --- | --- | --- | --- | --- | --- |
|  | Omron |  | ArteVu |  | Omron |  | ArteVu |  |
| H1 | 131 | 90 | 140 | 75 | 128 | 84 | 132 | 66 |
| H2 | 131 | 81 | 123 | 82 | 121 | 71 | 125 | 90 |
| H3 | 147 | 80 | 148 | 79 | 124 | 80 | 157 | 105 |
| H4 | 126 | 67 | 123 | 68 | 121 | 67 | 113 | 54 |
| H5 | 103 | 67 | 94 | 62 | 95 | 62 | 94 | 65 |
| H6 | 95 | 60 | 94 | 57 | 97 | 60 | 88 | 57 |
| H7 | 121 | 76 | 131 | 74 | 123 | 80 | 137 | 77 |

| Winter<br>ID | After exercise |  |  |  | After 2 minutes |  |  |  |
| --- | --- | --- | --- | --- | --- | --- | --- | --- |
|  | Omron |  | ArteVu |  | Omron |  | ArteVu |  |
| H1 | 89 | 57 | 85 | 55 | 95 | 62 | 89 | 55 |
| H2 | 104 | 73 | 112 | 72 | 102 | 71 | 112 | 72 |
| H3 | 146 | 98 | 152 | 82 | 151 | 89 | 149 | 81 |
| H4 | 137 | 80 | 132 | 70 | 133 | 76 | 127 | 65 |
| H5 | 127 | 81 | 130 | 81 | 125 | 80 | 130 | 80 |
| H6 | 89 | 57 | 85 | 55 | 95 | 62 | 89 | 55 |
| H7 | 117 | 85 | 128 | 85 | 122 | 77 | 114 | 71 |

### Summer Performance

In the Solar Pole program, systolic BP (SBP) after exercise ranged from 92 to 111 mmHg (Omron) vs. 92 to 114 mmHg (ArteVu). In the Health and Wellness Classes, the postexercise SBP reached 175 mmHg (ID: H5) using the reference device, which the fingertip sensor captured at 176 mmHg.

### Winter Performance

Despite an average ambient temperature of 7.4°C, the correlation between devices remained robust. In the Solar Pole group, the mean difference in SBP immediately after exercise was minimal (Omron: 108.5 mmHg vs. ArteVu: 107.0 mmHg). In the Wellness class, the higher SBP levels after exercise (ID: H4: 165 mmHg; H5: 166 mmHg) were detected by the fingertip sensor (163 and 160 mmHg, respectively).

### Accuracy and agreement

Across all sessions, the fingertip-based monitor did not show systematic bias compared to the Omron cuff. The most remarkable finding was the device’s ability to maintain accuracy during the recovery phase (2 minutes after exercise), wherein BP typically decreases. In winter, despite potential peripheral vasoconstriction, the fingertip sensor’s readings closely mirrored the cuff-based readings.

### Three-way Analysis of Variance (ANOVA)

Three-way ANOVA was performed with SBP and diastolic BP (DBP) as the dependent variables and the season, time-of-day, and device factors as within factors.

#### 1. SBP

Three-way ANOVA was performed with SBP as the dependent variable and the season, time-of-day, and device factors as within factors. As presented in Table 3, no significant effects were observed for any factor. Furthermore, no significant interactions were observed between any of the factors (Table 3).

**Table 3.** Analysis of Systolic Blood Pressure.

| Effect | Factor | Results of Analysis of Variance |
| --- | --- | --- |
| Main effect | Seasonal factors | $F(1,10) = 0.002, p = 0.967$ |
| | Time factor | $F(1,10) = 4.151, p = 0.069$ |
| | Equipment factors | $F(1,10) = 0.053, p = 0.823$ |
| Interaction | Season × Time | $F(1,10) = 0.613, p = 0.452$ |
| | Season × Device | $F(1,10) = 1.065, p = 0.326$ |
| | Time × Device | $F(1,10) = 0.077, p = 0.788$ |
| | Season × Time × Device | $F(1,10) = 1.965, p = 0.192$ |

#### 2. DBP

Three-way ANOVA was performed with DBP as the dependent variable and the season, time-of-day, and device factors as within-subjects factors. As presented in Table 4, no significant effects were observed for any factor other than the device factor. Regarding the device factor, the Omron measurements showed significantly larger values than the AreteVu measurements. Furthermore, no significant interactions were observed between any of the factors (Table 4).

**Table 4.**
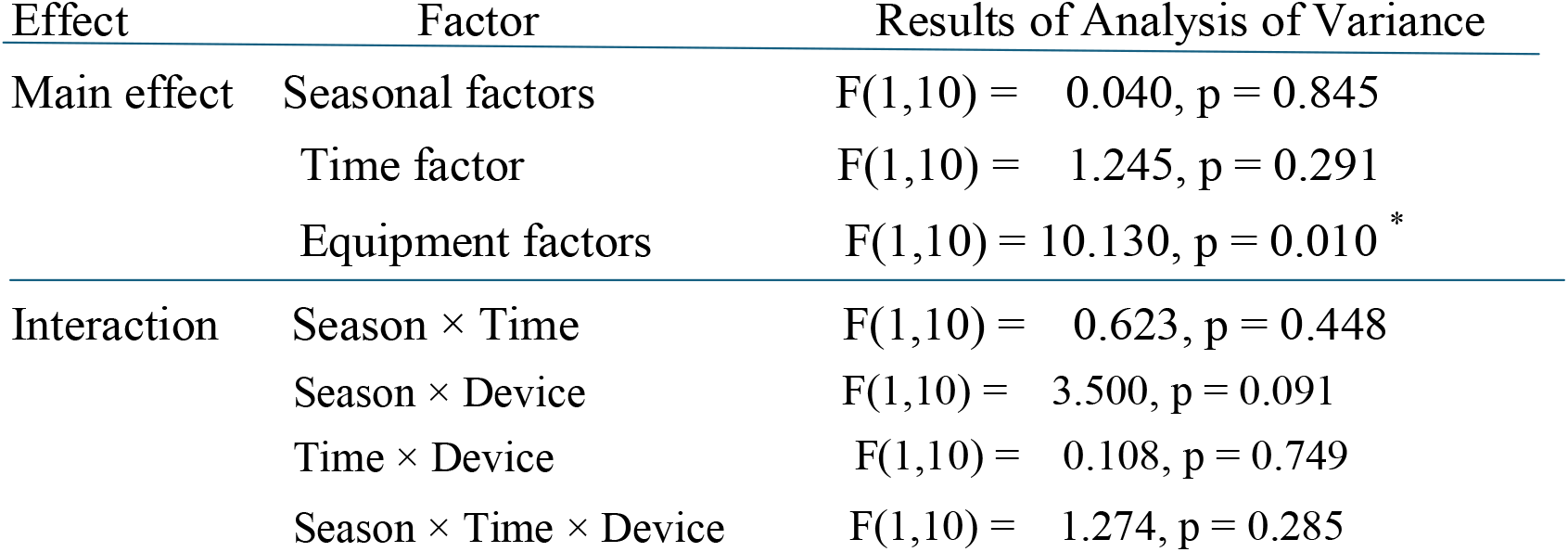
Analysis of variance for diastolic blood pressure measurements in both devices.

### Awareness Survey on BP Management

The poststudy questionnaire revealed a significant shift in the participants’ attitudes toward BP monitoring:

- **Ease of Use:** 90.9% of participants preferred the fingertip sensor due to the lack of arm constriction and simplicity of the measurement process.
- **Awareness:** 72.7% (8 out of 11) responded that using the continuous monitor increased their awareness of their own BP fluctuations. Several participants noted that seeing “real-time” fluctuations during exercise motivated them to be more proactive about their cardiovascular health.

## Discussion

### Principal findings

This study demonstrated the year-round utility of a cuffless fingertip-based continuous BP monitor, which may represent a potential paradigm shift in hypertensive management for older adults. The most remarkable finding is that despite the physiological challenges induced by winter, specifically cold-induced peripheral vasoconstriction, the fingertip sensor remained highly accurate and had a strong correlation (r = 0.88) with traditional cuff-based measurements. This finding confirms the robustness of fingertip-based technology for real-world application across all seasons.

### From “snapshot” cuffs to “continuous” ai management: a paradigm shift

Oscillometric upper-arm cuff devices have been the gold standard for home BP monitoring. Although reliable, these devices are limited to “snapshot” measurements that require the user to remain still and discontinue any physical activity, expose their arm, and endure cuff inflation. This physical and psychological burden has been a primary barrier to long-term adherence.

Our findings indicate that fingertip-based continuous monitoring is not merely a supplement to cuff-based BP monitoring but a **superior alternative** that could eventually replace traditional BP monitoring. BP management may enter a new era by integrating this technology with smartphone connectivity and AI-driven analysis. Additionally, continuous data streams allow AI to detect subtle fluctuations and early signs of cardiovascular risk that “snapshot” measurements would inevitably miss. This transition from discrete, manual tracking to automated, continuous, and intelligent monitoring represents a fundamental evolution in preventive medicine [11][12].

### Physiological robustness in winter

The increase in variance noted during winter (3.1 ± 11.2 mmHg for SBP) likely reflects the complex interplay between the environmental temperature and exercise-induced sympathetic activation. However, as the device’s readings remained within clinically acceptable limits, this confirms that the underlying algorithms effectively compensated for seasonal peripheral vascular changes. This robustness is critessentialical for an “always-on” AI monitoring system to function reliably in diverse real-world environments.

### Empowering older adults through digital health

The 72.7% increase in BP awareness among participants highlights the transformative power of usability. Eliminating the BP cuff removes the “friction” in BPmonitoring, transforming a burdensome task into a seamless routine of a healthy lifestyle. When combined with upcoming smaller and more portable models, this device may empower older adults to proactively take control of their health through real-time feedback and AI-mediated insights.

## Conclusion

In conclusion, this study provides evidence that fingertip-based continuous BP monitoring is a feasible method for year-round BP monitoring. As we transition toward cuffless BP monitoring and AI-integrated health systems, the traditional reliance on manual cuff inflation will likely diminish. This study also provides the essential evidence required for the widespread adoption of next-generation continuous BP monitoring devices, paving the way for a proactive and data-driven lifestyle management that may prevent cardiovascular events.

## Data Availability

All the data is included in the tables in the paper.

## Conflict of interest

The ArteVu blood pressure monitor used in this study was loaned free of charge by CARDIO RING. However, the study was conducted impartially.

## Notes

### Competing Interest Statement

The authors have declared no competing interest.

### Funding Statement

The author(s) received no specific funding for this work.

### Author Declarations

This study was approved by the Ethics Committee of Kyorin University (Approval No. 2024-28 and 2024-72).

## References

1. World Health Organization. Technical specifications for automated non-invasive blood pressure monitoring devices with cuff. Geneva: WHO; 2020.

2. Kario K. Management of hypertension in the digital health era: small devices for big data. Hypertension. 2018; 72:531–539.

3. Hoshide S, Kario K. The current status and future of home blood pressure monitoring. Hypertension Res. 2023; 46:1–10.

4. Trivedi R, Chapman N, Mishra SR, Nelson MR, Chow CK, et al. Attention to blood pressure cuff sizes is important for home and in-clinic blood pressure measurement. Hypertension Research (2025) 48:2719–2724.

5. Alperovitch A, Lacombe JM, Hanon O, Dartigues JF, Ritchie K et al. Relationship between blood pressure and outdoor temperature in a large sample of elderly individuals. Arch Intern Med. 2009; 169:75–80.

6. Mizutani N, Enomoto Y, Okamoto H, Nishizawa S, Murakami S, et al. Validation of a fingertip continuous blood pressure monitor and an awareness survey on blood pressure measurement. Cardiol Res Rep. 2025;7.

7. Stergiou GS, Mukkamala R, Alberto A, Kyriakoulis KG, Mieke S, Murray A, et al. Cuffless blood pressure measuring devices: review and statement by the European Society of Hypertension. J Hypertension. 2022; 40:1449–1460.

8. Omron Healthcare. HCR-7204T instruction manual and technical validation.

9. Bland JM, Altman DG. Statistical methods for assessing agreement between two methods of clinical measurement. Lancet. 1986; 1:307–310.

10. Pescatello LS, Buchner DM, Jakicic JM, Powell KE, Kraus WE, Bloodgood B, et al. Physical activity to prevent and treat hypertension: a progressive narrative review. Curr Hypertens Rep. 2019; 21:87.

11. Matsumura K, Rolfe P, Toda S, Yamakoshi T. Cuffless blood pressure estimation using only a smartphone onitor. SCIENTIFIC Reports (2018) 8:7298 DOI:10.1038/s41598-018-25681-5

12. Cornelissen VA, Smart NA. Exercise training for blood pressure management in adults with hypertension. Am J Hypertens. 2013; 26:1457–1467.

13. Wildenbos GA, Jaspers M.W.M, Schijven M.P, Dusseljee-Peute L.W. Mobile health for older adults: Using an aging barriers framework to classify usability problems. International Journal of Medical Informatics 124 (2019) 68–77

14. Cao W, Miks MW, Liu X, Gregory ME, Addison D, Zhang P, et al. Effectiveness of mHealth interventions for self-management of hypertension: systematic review. JMIR mHealth uHealth. 2022. 16–20.

15. Kario K. Essential manual on perfect 24-hour blood pressure management. New York: Wiley-Blackwell; 2018.

16. Mancia G, Kreutz R, Brunstrom M, Burnier M, Grassi G, Januszewicz A, et al. 2023 ESH guidelines for the management of arterial hypertension. J Hypertens. 2023; 41:1874–2071.

17. Stergiou GS, Alpert B, Mieke S, Asmar R, Atkins N, Eckert S, et al. A universal standard for the validation of blood pressure monitoring devices: Association for the Advancement of Medical Instrumentation/European Society of Hypertension/International Organization for Standardization (AAMI/ESH/ISO) collaboration statement. Hypertension. 2018; 71:368–374.

